# Adolescent idiopathic scoliosis (AIS) non-operative treatment in HUCSK of Kosova- a 7 month study

**DOI:** 10.1101/2020.08.11.20172627

**Authors:** Gresa Lokaj

## Abstract

I.

**Introduction:** Scoliosis is defined as a deviation from midline in the frontal plane, and rotation of the apex of the curve from ≥10° in AP radiography. Treatment of scoliosis is based in type of scoliosis, region of scoliotic curve, curve magnitude, bone maturity, gender, risk of scoliotic curve progression, other symptoms like and patient’s opinion about his spine shape. Treatment choices of AIS are observation, physical therapy, corsets and surgical treatment.

**Purpose:** Research of patients with AIS focusing in its characteristics, clinical presentation, diagnosis and a detalized research in non-operative treatment of AIS focusing in treatment choices and criteria of treatment.

**Material and methods:** The study is based in retrospective research September 2018-March 2019 (7 months) in HUCSK- Specialized outpatient clinics of Orthopaedics. Data is taken with special permission from Personal Data Protection

Office and Ethico-Professional Commity from specialized outpatient clinics of Orthopaedics system’s archive for patients with AIS of 10-18 years old.

**Results:** During September 2018-March 2019 period in specialized outpatient clinics of Orthopaedics-HUCSK, 250 cases with AIS of 10-18 years old with Cobb angle ≥10° are diagnosed and treated. The disease has a prevalence 1.40%. Based in gender women are more affected than men in a ratio 2.01:1. Most common form of AIS is the one that affects thoraco-lumbal region of spine with 60.8% of cases. There’s found a correlation between scoliosis and kyphosis in higher levels of spine.About 89.7% of cases are light scoliotic curves according to Cobb angle and the female\male ratio increases at women with increasement of Cobb’s angle. Patients are treated with one or more forms of non-operative treatment: observation 10.4% of cases, observation and physical therapy 89.6% of cases and observation, physical treatment and corset TLSO 25.6% of cases. Cases that have undergone three forms of non-operative treatment are with Cobb angle ≥20°.

**Discussion:** The results of this study are supported from many studies made in Germany, Singapor, Grece, Turkey and USA, from the earlier and later years, with data exactly or closely to this study results.

## II. Introduction

Scoliosis is defined as a deviation of the normal vertical line of the spine consisting of lateral curvature with rotation of the vertebrae within the curvature. To be considered scoliosis, the angle created in the spine on postero-anterior radiography must be at least 10° and accompanied by vertebral rotation (1). Scoliosis is a deformity of the spine that involves anatomical structures such as bones, muscles, ligaments and organs. In essence, scoliosis is an irreducible and progressive 3-dimensional deformity in contrast to the scoliotic attitudes, which is only a compensatory deviation in the frontal plane to reduce the asymmetry of the foot or secondary deformities from infections, osteomas, vertebral disc prolapse, etc. (6). Any curvature in the frontal plane is considered scoliosis. In the sagittal plane there are two lordotic physiological curves: in the cervical and lumbar region as well as two kifotic curves in the thoracic and sacral region. Increasing or decreasing the angle of these curves is considered pathological and of clinical significance (3). Scoliosis involves asymmetry of the spine and thoracic cage in all three planes of the body: frontal, sagittal and transverse (4).

**Fig 1.**
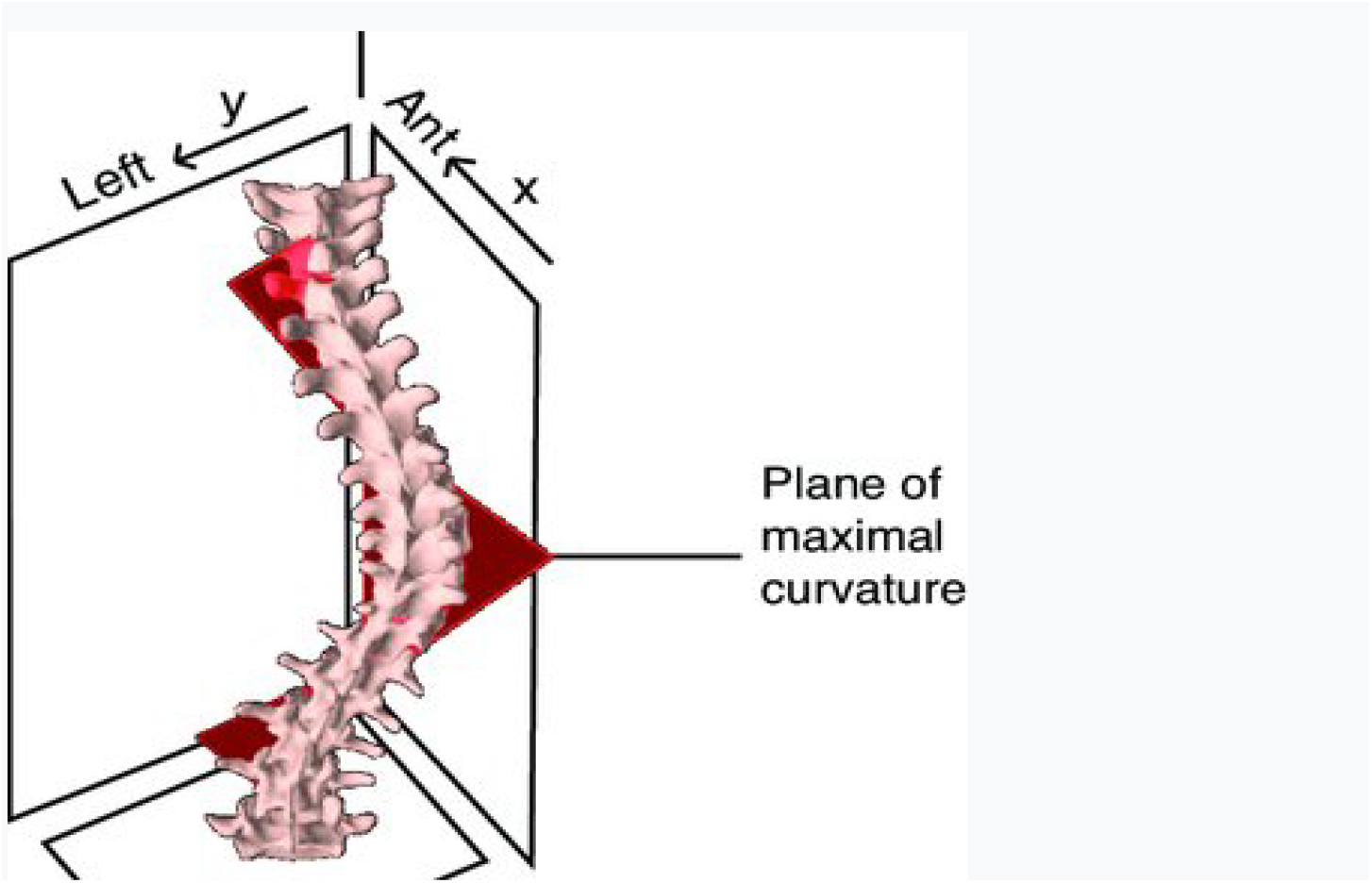
Maximal plan of curvature (Spine volume 39. (2014). *Lippincot Williams & Wilkins*. 10. pp. 601-606).

The causes of scoliosis vary and are mainly classified as: congenital, neuromuscular, syndromic, idiopathic or as a secondary curvature of the spine. The most common form encountered in practice is the idiopathic form (1). Scoliosis can develop in infants or early childhood. The usual age of onset of scoliotic curvature is 10-15 years, appearing the same in both sexes, but the progression of curvature size is 8 times higher in females than in males (5). The development of the disease is related to the etiology and age at which the scoliotic curvature appeared and together with the patient’s medical history, physical examination and radiographs taken of the spine determine the method of treatment (1).

### 2.1. Epidemiology of the disease

According to 2015 data, scoliosis is considered a major global problem affecting 28 million people, most of whom are children aged 10-16. This problem indirectly affects parents, family, partner, etc. who take care of their relatives with scoliosis and who are around 100 million people. Meanwhile, it is expected that in the coming years the number of these patients will increase dramatically. According to this study the number of patients will increase every year for 250,000 new patients and sometime in 2050 there will be approximately 36 million patients with scoliosis (9). In about 85% of cases, scoliosis is idiopathic (10). If the degree of deformity is ≥10° then the female / male ratio is 1.4: 1 and if the degree of deformity is more than 30° then the female / male ratio is 5: 1 (11). Adolescent idiopathic scoliosis develops at the age of 11-18 years and accounts for 90% of all idiopathic scoliosis (13). The prevalence of adolescent idiopathic scoliosis is 0.47% -5.2% and usually affects more women than men 1.5: 1 to 3: 1 and this ratio increases in favor of women with increasing age. About 90% of them will present with dextroconvex thoracic curvature (12). According to a 2010 meta-analysis, about 38% of children in schools who underwent screening tested positive for adolescent idiopathic scoliosis and were referred to an orthopedist (17).

### 2.3. Classification of scoliosis

Scoliosis is divided into two major groups:

- non-structural (functional); this type of scoliosis is a temporary (reversible) condition where the spine is normal and the deformity results from another problem. This group includes: postural scoliosis, hysterical scoliosis, scoliosis due to irritation of nerve roots, discrepancies in the length of the lower extremities, shortening of the lower extremity, infections and inflammations of the abdominal region (eg appendicitis) or spine.
- struktural; in this group of scoliosis the spine is not anatomically normal and the deformity is caused by the influence of various internal or external factors or by another disease and are usually irreversible. This group includes scoliosis: idiopathic, congenital, neuromuscular, neurofibromatosis, rheumatic diseases, trauma, contractures, osteochondrodystrophy, bone infections, metabolic disorders, tumors, mesenchymal disorders (Ehfan-Marfan syndrome, etc.). (10, 14).

Idiopathic scoliosis, which accounts for 85% of all scoliosis, is divided into:

- Infantile: appears from birth to the age of 3 years (0-3 years). It usually resolves spontaneously in 80% of cases and in 20% of cases the scoliosis progresses and requires further treatment.
- Juvenile: appears from the age of 4 to 9 years;
- Adolescents: appears from the age of 10 until bone maturity (14, 2).

Another classification of idiopathic scoliosis is based on the age of the child at the onset of the disease and is divided into:

- early onset, age 5-7 years;
- late onset, age from 7 years to maturity (12).

### 2.4. Etiopathogenesis of the disease

The etiology of idiopathic scoliosis is still unknown and multifactorial (2). The pathogenesis of juvenile idiopathic scoliosis includes: an initial stage in which a small curvature develops due to a defect in the control of the neuromuscular system, in the second stage during growth in adolescence where the curvature is exacerbated by biomechanical factors and further neurological dysfunction contributes to progression of curvature (25). Based on the fact whether the abnormality directly or indirectly affects the growth mechanisms of the spine, the authors have divided these factors into two groups:

#### 2.4.1 Internal factors

genetic, biomechanical factors of erectile posture, asymmetric growth of the spine, abnormalities of discs and ligaments (15)

Genetic factors - epidemiological studies have shown that there is a polymorphism of a nucleotide at different loci of the chromosome giving dysfunctions such as: structural abnormalities of connective tissue, disorder of calcium and bone metabolism and dysfunction of growth hormone and signaling factors (56). Many studies have found an increase in the incidence of scoliosis in families with members affected by scoliosis, thus proving links between the genetic component and the etiology of scoliosis, such as the Risenborough and Wynne-Davies study where they found scoliosis in 11.1% of relatives first instance, the study of Kesling and Reinker who confirmed 73% concordance of scoliosis in 37 monozygotic twins in which 27 of the twins were both affected with scoliosis and of the 31 dizygotic twins 36% were affected. Then fingerprints also confirmed the association and found concordance of idiopathic scoliosis 92% in monozygotes and 63% in dyzigots. The exact mode of inheritance of scoliosis has not been found except in some families where it appears as autosomal dominant, X-linked, recessive but some genes have been found that are associated with the occurrence of scoliosis, such as: SNTG1 (gamma-1-syntrophin) with location 8q11.22, ESR1 (estrogen receptor-1) located at 6q25.1, CHD7 (chromodin helicase DNA binding protein 7) located at 8q12.1 (19).

Spinal cord growth abnormalities - the development and progression of scoliosis is related to the time of rapid growth in adolescence where different ratios of right and left spinal cord growth give asymmetry and emphasize the asymmetric biomechanical load as well as the Heuter Volkman effect. The Heuter Volkman effect is created by the law of the same name according to which bone growth is slowed by increased compression and distraction, and under the influence of gravity an initiated structural asymmetry will give stress to the concave area and produce increased curvature. Biomechanical studies have shown that the affected vertebrae vary: their curvature, length, and internal load (39). And in this way a vicious cycle will be formed in favor of the progression of scoliosis. According to Dickson the anterior growth of the spine leaves aside the posterior growth producing hypoxia of the spine and affecting the rotational deformity of scoliosis. But this theory is taken more as secondary than as primary. Yet another theory speaks of abnormal centers of joint development or so-called Schmorl cartilages which consist of separate and symmetrical synchondroses that join the vertebral body in each semiposterior arch and function normally symmetrically. This theory is also supported by MRI observations of children with scoliosis in which it is observed that the vertebral pedicures are asymmetric and by the fact that aggravated idiopathic scoliosis is synchronous with the time in which these centers are active. The development of scoliosis is also influenced by high levels of growth hormones and the characteristic morphometry of the body. Many studies show that adolescents with scoliosis are taller than their peers.

Intervertebral disc abnormalities - recent studies speak of collagen abnormalities of the intervertebral discs in patients with scoliosis. Montanaro found that polymorphism in a single nucleotide of Matrilin-1 (MATN1-protein which ensures the distribution of chondrocytes in the growth plane) affects the disorder of chondrocyte distribution and results in scoliosis, whereas polymorphism in only one nucleotide of the receptor sciatica nucleotide idiopathic (18).

#### 2.4.2 External factors

asymmetry of the body, abnormalities of the nervous system, abnormalities of the muscles or ribs of the thoracic cage, osteoporosis, abnormalities of platelets or melatonin (15)

Asymmetry of the body- according to careful observations of curvatures of the spine during the growth phase in persons without scoliosis with increasing age a greater curvature is observed on the right side of the dorsal part of the spine. In addition, the initial source of asymmetry is thought to be visceral rotation during the embryonic stage.

Connective tissue abnormalities - because scoliosis is common in diseases such as osteogenesis imperfecta, Marfan syndrome and Ehler-Danlos syndrome, these patients have been observed and the presence of abnormalities in the elastic fibers and yellow ligaments has been detected. However, the veracity of this study casts doubt on whether these connective tissue abnormalities are causes or consequences of scoliosis (15).

The role of spinal musculature - asymmetry of the erector spinae muscle since the XIX century has been thought to be the cause of scoliosis for this reason and the transverse spinal muscle tenotomy has been practiced as a form of treatment. However, even here we have the doubt whether this asymmetry comes as a result of scoliosis or is its cause. Histological analysis of paravertebral muscle has found denervative changes, ultrastructural changes in the sacrolema of myotendinous ligament supporting a new concept in the etiopathogenesis of scoliosis, that of primary muscle disorder (35).

Hormonal factors - girls with scoliosis have lower body mass than other girls of the same age and this is attributed to the psychological difficulties associated with deformity (23). Recent studies, however, have suggested that scoliosis may be due to metabolic and endocrine diseases that are specific to a morphological body type. This is observed in patients treated with growth hormone but not in all scoliotic patients changes in the concentration of this hormone in the blood are observed. Delayed puberty affects longer exposure to deforming stress from erectile posture which contributes to the onset of scoliosis. According to a study by Lebouf and colleagues, estrogen has an impact on the development of idiopathic scoliosis but not on the degree of curvature, while later it was discovered that the impact lies in the insufficiency of estrogen receptors which in addition to appearance also play a role in curvature of the spinal cord (20). The low bone density in young girls speaks of disorders in estrogen which gives osteopenia and as a result can give deformities of the bone matrix. According to a 2016 study, osteopenia was found in 30% of patients with adolescent idiopathic scoliosis but it has not yet been established whether it is a cause or consequence of scoliosis (21). Related to this is the late age of onset of menarche which has a higher prevalence of juvenile idiopathic scoliosis (22).

The role of melatonin- according to a study by Moreau et al. the progressive character of a small scoliosis is associated with an abnormality in the melatonin receptor membrane (24). Melatonin receptors are located in the brainstem and in the gray matter of the dorsal part of the spinal cord, areas that are associated with postural control. Machida et al. found lower than normal serum concentrations in patients with progressive scoliosis as opposed to those with stable spinal curvature (16). The hormone melatonin also causes osteoblasts to grow and osteoclasts to shrink. Melatonin receptors are lower in patients with scoliosis thus giving fewer responses. The synergistic effect of 17-beta-estradiol and melatonin on osteoblasts in patients with scoliosis is higher than that of split (26). Polymorphism in the TPH-1 gene results in lower melatonin levels and thus in the development of idiopathic scoliosis (27).

The role of leptin and the autonomic nervous system- anomaly in the hypothalamus at leptin receptors may give asymmetry to the efferent pathways of the SNA which will further give trophic asymmetry involving the vertebral bodies.

Role of calmudoline - calmudoline is a protein that plays a role in muscle contraction and some authors have linked muscle tone disorders to calmudoline in the development of scoliosis (28). Meanwhile another study talks about the link between high levels of calmodulin in platelets and advanced stages of scoliosis. According to one hypothesis this anomaly could give microangiopathy at the level of the suppressed vertebral bodies producing asymmetric vertebral dystrophy (29). Higher levels of calmudoline have been found in convex lateral than concave muscles in adolescent idiopathic scoliosis (30).

Theories of the central nervous system - during the detection of spinal cord abnormalities or asymptomatic neurological syringes it has been observed by MRI that scoliosis may be the result of an attempt at protection by a short spinal cord (31). Roth proposed the theory of “uncoupled neuro-osseos growth” according to which there is a growing difference between the spine and the spinal cord, a theory that was also supported by MRI examinations (32). Whereas according to a 2011 study the length of the spinal cord and spine was significantly shorter in patients with juvenile idiopathic scoliosis than the control group (33). Idiopathic scoliosis is also associated with co-existing disorders such as brainstem asymmetry (60). Syringomyelia is associated with an increased incidence of scoliosis due to direct pressure on the sensory and motor tracts of the spinal cord. It is thought that abnormal circulation of spinal fluid also plays an important role in the development of scoliosis. Goldberg and colleagues observed cerebral cortex asymmetry in scoliotic patients (34). Meanwhile, according to the study of cerebral structures by MRI, abnormalities in the corpus callosum and stereotypical asymmetry of the internal capsule were observed in patients with scoliosis (36). Another theory speaks of disorders in the vestibular and balancing organ according to which Woody and colleagues found that children with hearing impairment have a lower incidence of developing scoliosis (37). Another reason for dysfunction of the vestibular and balancing organ is the presence of genetic and morphological abnormalities of the semicircular canals of the left inner ear, in which case the proprioceptive and postural organs are also affected (38). Most factors are still being studied whether they are the cause of adolescent idiopathic scoliosis or its consequence. However, there are many causes and factors that influence the occurrence, development or progression of idiopathic scoliosis and which are under study until a unified theory is reached regarding the etiopathogenesis of idiopathic scoliosis.

### 2.5. Clinical manifestation of the disease

Patients with juvenile idiopathic scoliosis are usually observed in clinical examination as body trunk asymmetry. This is also noticed by the patient or family members, during screening in schools or as a random finding during physical examination, chest radiography or any other medical imaging (40). Adolescent idiopathic scoliosis appears at the growth spurt where even the spinal cord is more predisposed to deform. The infantile form at 6-24 months of age, the juvenile form at 5-8 years of age and the adolescent form at the same time and the most common form at 11-14 years of age. While according to pubertal growth it corresponds to Tanner stages: S2, P2 in girls and S2, T2 in boys. The appearance of deformity in this period occurs due to the rapid and disproportionate growth of the extremities with the spine which stagnates slightly (60).

Adolescent idiopathic scoliosis is not usually associated with pain or neurological symptoms. Back pain depends more on the type of scoliotic curvature than on the degree of scoliotic curvature (47). However, in its early onset we have mild pain in the lower back which mostly comes from numerous activities which do not have sufficient support from the abdominal and back muscles or from the reduced flexibility of the lower back muscles. This mild pain is more manifested in the form of discomfort in moderately severe scoliosis and according to a study by Ramirez and colleagues only 23% of them complain of this symptom and 9% of them have had a pathological condition that explains it. Neurological symptoms, which are less common, come more as secondary from spinal curvature giving: weakness, sensory changes, disturbances in balance and gait, numbness, pain or even bladder and bowel control disorder (43). According to one study, body asymmetry can lead to psychosocial problems such as: lack of self-confidence, tendency to depression, suicidal thoughts and alcohol consumption (49). Rarely are symptoms that may come as a result of pressure on the organs of the heart or lungs, which are: shortness of breath or chest pain. The symptomatology depends on the type, size and magnitude of the curvature of the spine. The inspection shows asymmetry in the height of the shoulders where one shoulder looks higher than the other. There is an elevation or displacement of the body to the left or right when we have a primary curvature in the thoracic or lumbar part of the spine without a secondary curvature to compensate for it, resulting in asymmetry in the pelvic region with elevation one iliac crest more than the other which is accompanied by the appearance of one leg longer than the other. The most obvious sign is the prominence in the spine or the rib in the rib of the thoracic cage that comes secondarily from the rotational aspect of scoliosis (41).

**Fig. 2.**
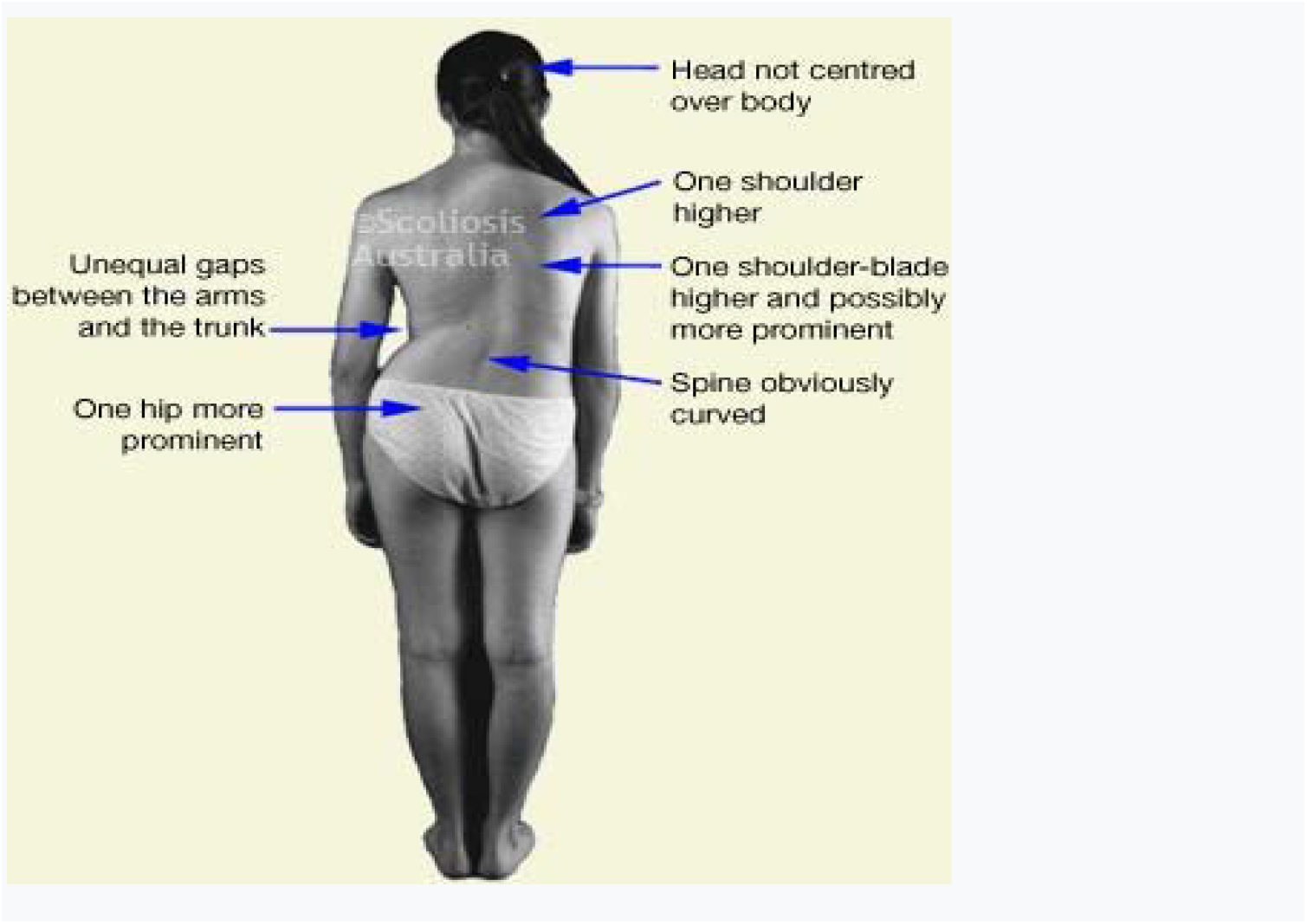
Figure shows signs of adolescent idiopathic scoliosis on inspection of a girl with right thoracolumbar scoliosis (Avaliable online: https://www.scoliosis-australia.org/scoliosis/about_scoliosis.html)

In addition, in the inspection of early scoliosis, deformity in the form of antiposition can be noticed in: clavicle, costal arches and deformity on the same side of the thoracic cage and in the back of the same side of the back, lumps are observed in the back and vice versa with the right side. As a result in women can be seen promotion of the breast of the same side forward. A slight turn of the head more on the opposite side of the curvature of the spine can also be noticed. The antiposition of the rib arches also affects the shortening of the extremity, a shortening which is relative and comes more due to the compensatory nature of the bend and the muscular tension in the extensor muscles of both sides of the back. On the opposite side with the shortened extremity is observed greater increase in the volume of the back muscles (M. erector spinae and M. latissimus dorsi). At the shortest extremity, a varus deformity can be seen in the lower third of the thigh and tibia, which affects the left side more, and as a result, one side of the back appears smaller than the other, and in scoliosis of the scales more severe in addition to the affected soft tissues is also observed hypoplasia of the pelvic bones. Further examination also shows asymmetry on the face, on the side of the curve, where: the ear, eyebrows, eye, nostril of the nose, the corner of the mouth and the sulcus nasolabialis are located lower than the other side of the curve opposite to the face (42).

### 2.6. Types and forms of scoliotic curves

There are different types and forms of scoliotic curves and that is why there are different classifications of them. For many decades great effort has been made to classify bends in the frontal plane to describe deformity, to predict progression, to implement an appropriate treatment plan, to determine the appropriate surgical strategy, and to define the biomechanical principles of corsets, and specific exercises for specific scoliotic curves (46). Initially the scoliotic curvature is defined by cranial vertebrae and caudal vertebrae bent mostly toward the concavity of the scoliotic curvature. The exact location of scoliosis is determined by the apex of the curvature. The apical vertebra is the vertebra that is most rotated in the scoliotic curve and is farthest from the vertical axis of the scoliotic patient. It is usually just one apical vertebra but there may be two. They have the most horizontal position. The caudal vertebra of the curvature defines the proximal and distal extension of a curvature where they are defined by the most horizontally displaced vertebra. The sacral centerline is the vertical line that symmetrically separates the sacrum and determines the balance of the spine relative to the pelvis (16).

**Fig. 3.**
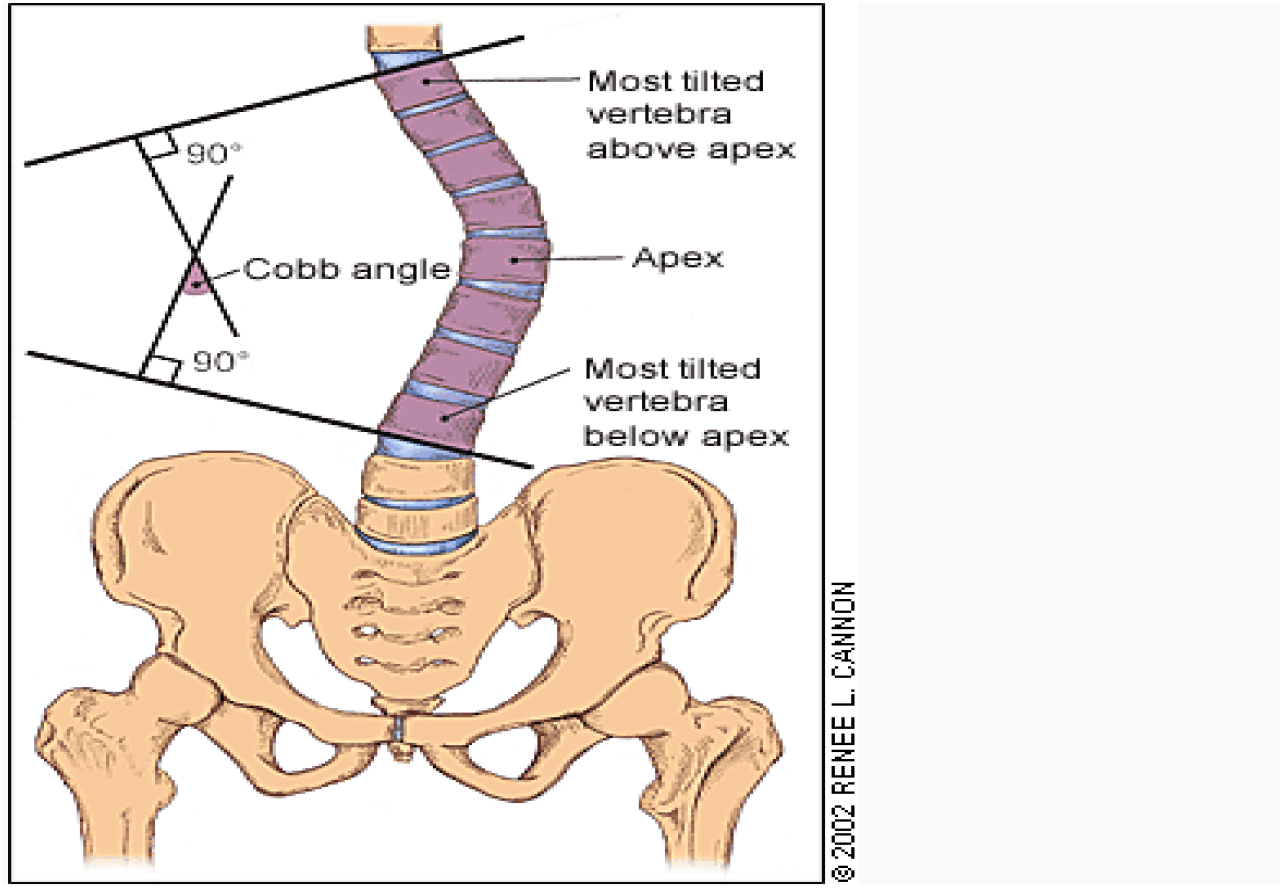
Cobb angle (Greiner KA. (2002). Adolescent idiopathic scoliosis: radiologic decision-making.).

One of the first classifications of scoliosis is that of 1948 by John Cobb. He was the first to describe major and minor, structural and non-structural curves and to prepare guidelines for their treatment (45). In 1950 Ponzett and Friedman presented their classification of scoliosis which described different approaches, progression and treatment for different anatomo-radiological forms of scoliosis. Moe and Kettleson classification which describes three curves: thoracic, thoraco-lumbar and lumbar, as well as 4 types of combined scoliotic curves: major thoracic / minor lumbar, double thoracic major / lumbar, double major thoracic / thoracolumbar and double major was among the most widely used classifications by orthopedists (46). Based on the flexion of the two most anatomically disordered vertebrae there are: dextroconvex and sinistroconvex curves (14). This type of division is important in distinguishing the types of scoliosis by age from James by 1954. Infantile idiopathic scoliosis in contrast to juvenile and adolescent scoliosis characterized by thoracic dextroconvex curvature, these are characterized by thoracic sinistroconvex curvature. Adolescent idiopathic scoliosis where girls are most affected is characterized in 80% of cases by dextroconvex thoracic curvature and lumbar synistroconvex (50). The SRS (Scoliosis Research Society) has made a descriptive technical classification depending on the position of the scoliotic curvature as well as the apical vertebrae (16).

Scoliosis usually has two curves: the primary curve and the secondary curve. The primary curvature is the first idiopathic scoliotic structural curvature that occurs due to various etiological factors of scoliosis while the secondary curvature is the compensatory curvature which enables the achievement of balance which is disturbed by the primary curvature (14). This is the basis of King’s 1983 classification of scoliotic curves, and we are dealing with the division of curves into 5 groups depending on the location of the curve (44).

This classification system was based on Moe’s experiences in the surgical treatment of AIS patients according to the Harrington method, and to this day is still used to design corsets for the treatment of scoliotic curvature. The disadvantage of this system is that it does not provide accurate inter and intra-observational data to determine the treatment protocols and the exact level of the spine for fusion in operative cases. This shortcoming led to the creation of Lenke’s new classification system in 2001 according to which surgical treatment is significantly facilitated even though 15% of surgeons still deviate from the algorithm in fusion-level selection. This classification has failed to include the rotating element and is therefore not used in non-operative treatment with corsets. It is less complicated than the classification according to King. The Lenke classification includes 6 types of curves, 3 lumbar modifiers and 3 sagittal thoracic modifiers. It is a 2D classification system and all types of curves can be included in this classification (48).

Regarding the division of scoliosis for treatment with corsets has historically been based on the difference between single or double curvature. The corset separation system according to Providence has 3 basic models: lumbar, thoracolumbar and double curvature with a possible extension for thoracic curvature. Lehnert-Schroth further refined this system by using it for physical therapy as well. This system included triple and quadruple curvature models to distinguish single thoracic curvatures with little or no lumbar curvature from true double curvatures accompanied by lumbo-sacral compensatory curvature. Cheneau later defined corset patterns as “triple curved scoliosis” as a single scoliotic curve and “quadruple curved scoliosis” as double scoliotic curves (46). There is a classification of scoliosis according to SOSORT protocols of 2016 that is used nowadays.

Researchers have recently provided stereoradiographic measurements of spinal deformities, in which case the deformity is seen in the 3D plane (45).

### 2.7. Diagnosis of the disease

An important part of diagnosing scoliosis is the patient history (50). Adolescent idiopathic scoliosis presents as spinal and lumbar (waist) deformity with atypical back pain and usually complains that the blouses do not fit the body well, the body trunk lateralizes, and the arm rubs ipsilaterally with the pelvis (51). The points at which the doctor should focus on the diagnosis of scoliosis are: age when the deformity is observed, the patient’s maturity, the presence of back pain, neurological symptoms (gait disorders, weakness or sensory changes), the patient’s preoccupation with appearance and shape of the spine and family history (52). Family history plays an important role in diagnosis as the presence of idiopathic scoliosis in the immediate family increases the prevalence of the disease by 7 times if it has a sibling and 3 times if it has one parent. The extent of pubertal physical changes and menarche are also analyzed (16). In the case of newly diagnosed scoliosis the emphasis falls on the patient’s current or potential illnesses. If the patient has back pain especially at night the suspicion falls on the tumor, changes in bowel or bladder function speak to neurological disorders (50). The triad that identifies a non-idiopathic scoliosis is the age at onset of the disease, the degree of curvature progression, and the presence of neurological symptoms.

Physical examination in women but also men is done in swimsuits or underwear to better notice the deformity and other physical changes (16). Physical examination of the scoliotic patient should begin with general appearance, skin, neuromuscular system before assessing the shape of the spine. The points to focus on in the physical examination are: measurement of body height, analysis of the patient’s gait, shape of the foot, skin inspection, assessment of pubertal development (Tanner scale), complete neurological examination, symmetry of shoulders and iliac crests as well as Forward bending test. The rapid longitudinal growth of the body raises suspicions of Marfan or Ehler-Danlos syndrome, pes cavus accompanies Charcot-Marie-Tooth type diseases, skin pigmentation like café au lait speaks of neurofibromatosis and lower back hairs for spinal dysraphy. During the inspection, attention is paid to the symmetry of the shoulders and iliac ridges, the position of the scapular blades, the shape of the waist and the discrepancy in the length of the lower extremities viewed from front and back while the patient is standing. The elevation of one side of the pelvis is also determined by palpation of the iliac crest and spina iliaca inferior posterior bilaterally while the patient is standing and has his legs fully extended. The measurement of the trunk deviation is also done by means of the pendulum (“bullet bob”) which is released from C7 to the sacral region, thus being measured in centimeters of deviation. At the end of the physical examination, a forward bending test is performed, which is performed by bending the body forward, keeping the knees straight and the palms of the hands together, during which time the examiner looks for any asymmetry along the contour of the spine. Examiner looks from the back for lower trunk rotation, from the front for upper trunk rotation and laterally for eventual kyphosis. If we have prominence on either side of any region of the spine: upper thoracic, middle thoracic, thoraco-lumbar, and lumbar then the test is positive. The height of the prominence is measured in centimeters and the prominence in general, with a scoliometer. A trunk rotation magnitude with this test of 5-7° on RTG indicates a Cobb angle of 15-20° (52, 16). For the diagnosis of thoracic curves there is a sensitivity of 92% and a specificity of 60% (53). This test is also used to distinguish the scoliotic attitude from the real scoliosis where in the first the test is negative (14). Once the test is positive, it is continued with RTG postero-anterior, antero-posterior, lateral to the leg and lateral flexion of the spine, which are standard for the diagnosis of scoliosis. In addition to the spine, the clavicle and iliac crest should be included in the radiograph to obtain an ideal RTG (51). In infantile patients and those with severe neuromuscular involvement RTG can be performed in a sitting position (16). Lower limb discrepancies should be corrected with any body placed below the shorter leg before RTG is performed (54). Attention is paid to both pedicures of each vertebra in the RTG. In addition to the deformity in the coronal plane, the diagnosis is confirmed by the rotation of the spine with the apex of curvature representing the most rotated vertebra (52).

To measure the degree of curvature the Cobb angle is used where in the made RTG the straight line is drawn along the upper part of the rotating cranial vertebra and the lower part of the caudal vertebra and the verticals drawn on these two lines by their union form the angle which if it is 10° or more and has the rotating element speaks of scoliosis. Bends between 0 and 10° are not considered true scoliosis

**Fig. 4.**
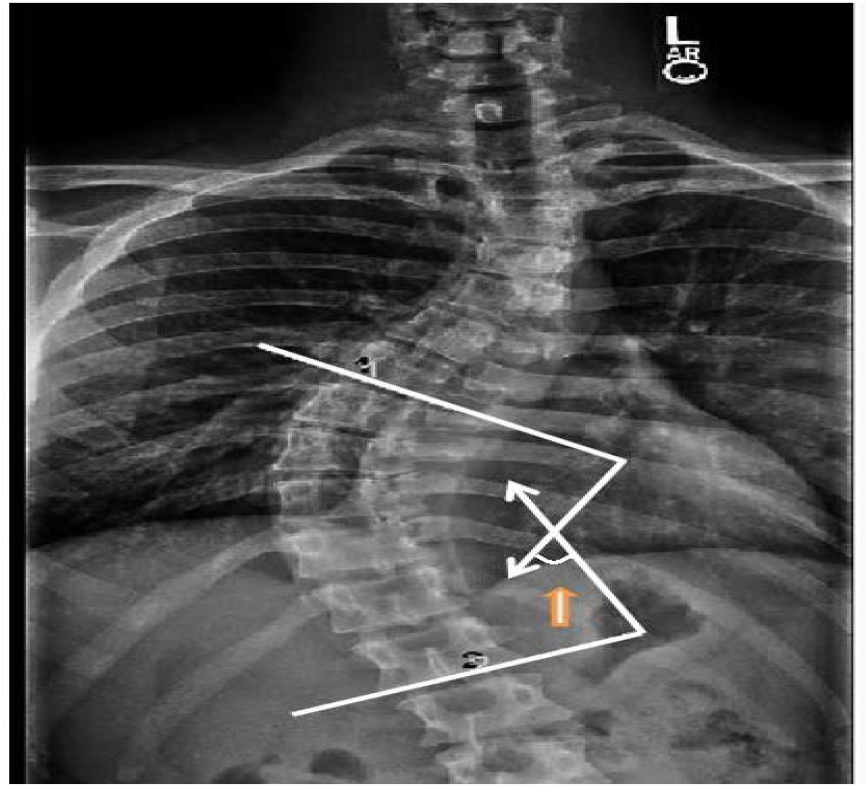
Measurement of Cobb angle (Avaliable online: http://xrayhead.com/measure/show_measurement.php?i=40).

According to Cobb’s angle scoliosis is divided (51): mild curvature 10-30°, moderate curvature 31-50° and severe curvature >50°.

The number of RTGs depends on the age and progression of the scoliotic patient curvature but never more than 3 RTGs are performed (14). If we have more than one bend, the Cobb angle measurement is made for each bend. The limited number of RTGs and RTGs done postero-anteriorly are preferred to reduce the chance of developing breast and thyroid cancer as the prevalence increased 1-2% during its treatment (16). If there is no curvature in the transverse plane (rotational) then there is talk of other pathologies such as: tumors, spondylosis, spondylolisthesis, infection, etc. (52).

Further examination based on back pain includes: CT, bone scintigraphy and MRI which are not part of the routine examination of scoliosis but serve to rule out other diseases. MRI is used only in infantile patients, with less than 10 years in diagnosis, bone abnormalities, thoracic curvature, and abnormalities in neurological examination (52, 16). Once the disease has been diagnosed further research is done on the progression of the curvature. The classification according to Risser has high inter and intra-observational connectivity (51). The classic form for predicting the progression of scoliotic curvature is also the classic Risser sign from 1958 which observes the ossifying degree of iliac apophysis which is related to the state of bony maturation. It is a physiological form of measuring the maturity of the spine by comparing it with the ossification of the iliac apophysis and has values from 0-5, where 0 indicates that there is no ossification center yet and 5 ossification is complete. The smaller the Risser scale, the greater the chance of curvature progression (57). Another method of measuring bone maturity is the antero-posterior RTG of the wrist and left hand and is compared according to the atlas standards of Greulich and Pyle (59). In 2010 the ScoliScore AIS prognostic was developed designed for Caucasian men and women aged 9-13 years diagnosed with mild AIS (Cobb 10-25°) to help predict the progression of scoliotic curvature. This test is performed by means of the saliva where through the algorithm the results of the TaqMan genotype are converted with a numerical value of 1-200 and the higher the numerical value the greater the risk for progression of scoliotic curvature. The predictive safety of this test is 99% but the test does not predict the possibility of inheritance or the final progression of the curvature (55).

### 2.8. Treatment of the disease

Treatment of scoliosis is based on the type of scoliosis, scoliotic curvature region, curvature magnitude, bone maturity (age, menarche status, Risser scale), gender, risk of progression of scoliotic curvature, symptoms such as breathing problems and heart and the patient’s opinion regarding the shape of his back. Treatment options for AIS are observation, physical therapy, corsets, and surgical treatment. Considering other factors, the main goal is to keep the scoliotic curvature below 50° at maturity (52, 12). According to protocols approved by SOSORT in 2016 treatment and rehabilitation of idiopathic scoliosis during growth includes: methodology, background of idiopathic scoliosis, access of different patients to conservative treatment of scoliosis through practical algorithms and reviews, recommendations and evaluations physical, scoliosis-specific physical exercises (PSSE) and other conservative treatments. About 10% of those diagnosed with idiopathic scoliosis undergo conservative treatment and 0.1-0.3% undergo operative correction (60). According to some epidemiological studies in adolescents newly diagnosed with scoliosis 10-15% of cases have progression of curvature and 22-27% of cases have spontaneous improvement of curvature (56). The main purpose of conservative treatment is to stop the process of progression of scoliotic curvature and to avoid surgical treatment. Other goals include improving lung function, treating pain, improving aesthetic appearance, postural balance, and reducing psychological distress (58). Adolescent idiopathic scoliosis is a benign process which has no increased mortality.

#### 2.8.1 Observation

Since most patients have idiopathic scoliosis with an angle of less than 20° and only a small number of them are treated, most patients are only observed (16). Slight scoliotic curves are observed and followed by RTG and clinical examinations every 6-12 months until bone maturation or closure of the epiphyses and physical therapy is performed to strengthen the back muscles and relieve pain (51). Scoliotic lesions appearing at the peak of growth are observed every 4-6 months with RTG (16). As technical assistance in observation, scoliometers are used to measure the degree of a lump in the ribs and lumbar swelling, as well as stereoradiography which shows the complete shape of the spine by means of rays (2).

#### 2.8.2 Physical therapy

begins with mild scoliotic curves and the most appropriate form of therapy is Schroth therapy. This physical therapy has been shown to be successful in reducing the progression rate of scoliotic curvature or in stopping the progression of several weeks of treatment compared to patients who have not undergone physical therapy at all (2). Electrical stimulation or TENS also plays an important role (16). In addition physical therapy maintains the flexibility of the spine and reduces the description of corsets (58). In the physical treatment of scoliosis PSSE (Physiotherapy Specific Scoliosis Exersices) is used which is effective in scoliotic patients with mild to moderate curvature in improving Cobb angle, spinal rotation angle, pain and quality of life (61).

#### 2.8.3 Treatment with corsets

Standards for the treatment of idiopathic corset scoliosis are grouped into 6 domains: experience / competence, behavior, description, construction, corset control, and follow-up (60). The corset is designed to exert external force on the spine during the growth phase in adolescence to prevent progression (64). According to a study, treatment with corsets enables the prevention of severe scoliotic curvature 20-40° in 72% of adolescents while in 28% progression over 50° thus undergoing surgical treatment. In the group without treatment with corsets, the rate of curvature progression reached 52% (56). Accepted indications for treatment with corsets are: bends between 25° and 35° with proven progression 5°< bends 30-40°, immature patients who have at least one year to complete growth, Risser <3 and the patient, eventually the motivated parent (62). Medium Cobb angle bends 30°> and Risser scale 0 or 1, heavy Cobb angle curves 30-45° and Risser scale 2 or 3 and in bends with an angle of 20-30° with progression over 5° in 6 months corsets are used as a supportive measure in treatment and not as a definitive treatment (2, 51). There are two conditions for corset treatment to be successful: suitability and primary corset treatment. The duration of wearing the corset during the day also plays a role: the longer the better the results, the regular wearing of the corset and the support from the family (58). A Rowe study with associates has shown that wearing a corset for 23 hours is successful in 92% of cases (12). According to a meta-analysis by the SRSP (Scoliosis Research Society Prevalence) and the NHC (Natural History Comitte), wearing a corset for longer periods of time provides correction and inhibition of the progression of the most effective scoliotic curvature (63). Most corsets should be worn 16 hours a day for 2-4 years or until bone maturation with the patient’s Risser scale 4-5 and every 6 months a standing RTG should be done to monitor the effect of the corsets. When corset placement is indicated, RTGs are initially performed prior to corset placement so that these RTGs are compared with subsequent RTGs and treatment efficacy is monitored. RTGs are also performed with corsets worn in order to observe the frontal and lateral balance of the spine and the degree of improvement of the scoliotic curvature. This improvement should be 30-70% for the treatment to be permanently effective after bone maturation. Early placement of corsets can regress scoliotic curvature under certain conditions as in premenstrual women with good corset fit. To track curvature improvement some prefer RTG with and without corsets but RTG without corsets is done 24-48 hours after the corset is removed to enable an adaptation period (51). Discontinuation of corset wearing occurs after the end of growth, Risser rate 5 and 2 years after menarche (62).

There are different types of corsets but the most commonly used are thoraco-lumbar corsets, namely TLSO (thoraco-lumbo-sacral orthosis) and CTLSO (cervico-lumbo-sacral orthosis). The Milwaukee corset is CTLSO which is used for thoracic and double scoliotic curves. Makes active correction with neck support enabling longitudinal traction between the skull and pelvis and lateral translational force through the cords connected to the chest, but is used less frequently due to large immobilization. The underarm corsets are the Boston and Wilmington that have almost completely replaced the Milwaukee corset. The Wilmington Corset is a TLSO made of plastic and in the shape of a jacket, it is prescribed for 23 hours or 12-16 hours for scoliotic curves with less than 40°. Prefabricated orthoses like the Boston orthoses fit the back of the patient with special wadding and are used more in the US while orthoses ordered like the Chenau corset in Europe. Both correct scoliotic curvature through passive adaptation to adaptation. The Boston Corset is the TLSO used for all types of scoliosis and is for 23-hour wear. Along with the Milwaukee corset it is used as a superstructure in the treatment of thoracic curvature scoliosis with apex over T10

**Fig. 5.**
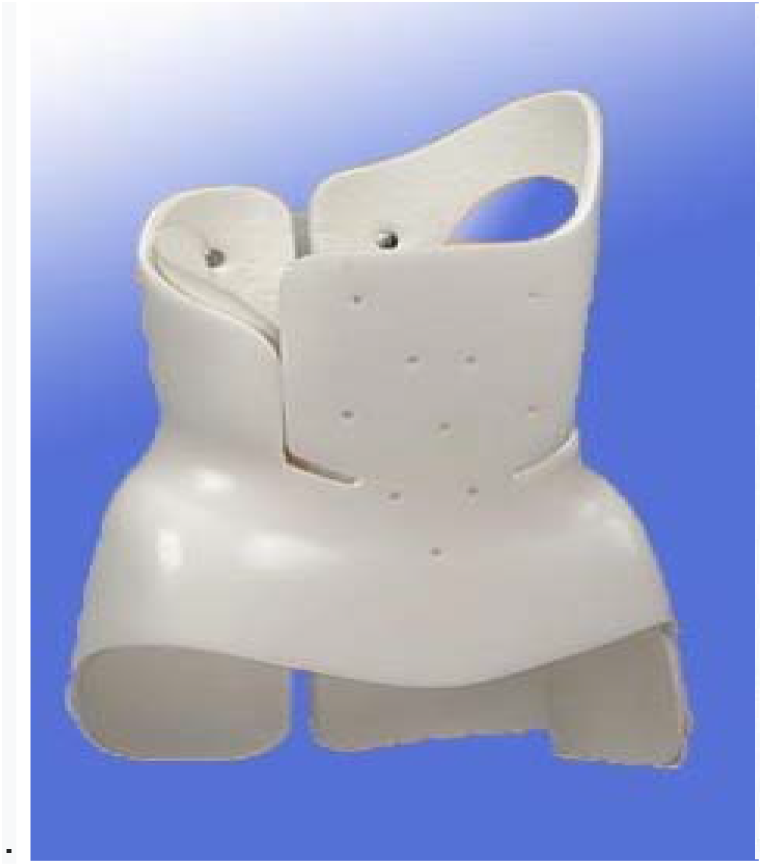
Cheneau Corset (Available online: http://www.bracingscoliosis.com).

**Fig. 6.**
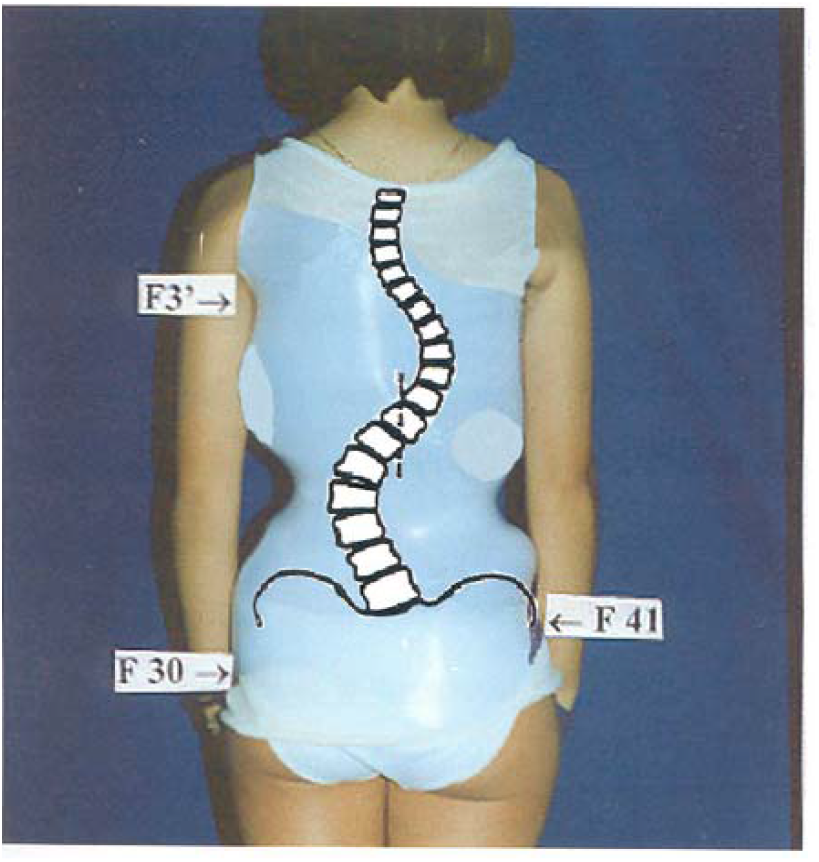
Boston Corset (Available online: http://www.britscoliosissoc.org.uk).

The Cheneau corset has a passive and active mechanism. The modified 3D Rigo-Cheneau corset is constructed in such a way as to exert force on the surface of the spine through ligatures that are connected and corrected in the best possible morphological form to light or medium juvenile scoliotic curves. Cheneau-light corset The Cheneau-Toulouse-Munster and Gresingen corsets are also variants of the Cheneau corset. The Lyon corset has a ligament extension mechanism at one point for 4 weeks and is then used overnight to ensure the visco-elasticity of the structure. The Spine-Cor dynamic corset uses specific corrective movement that depends on the type of scoliotic curvature and is constructed of elastic bands that provide neuromuscular and skeletal support. They are worn 20 hours a day for 18 months and their wearing is discontinued in Risser 4 or the end of adolescence. Night corsets include the Providence and Charleston corsets. The Providence Corset is constructed of an acrylic frame that exerts direct force on the scoliotic curve. Treats all single and double curves. The Charleston night corset requires a large flexion of the trunk impeding walking for this and is used at night and is preferred for single distal scoliotic curves like thoraco-lumbar and lumbar. Other types of corsets are also corsets: ART, Sibilla, Spinealite soft, Lapadula, Sforzesco, etc. Corset treatment of scoliosis in most cases is accompanied by physical therapy and observation, rarely used alone. Corset treatment failure is due to pain, corset maladaptation, heat, company, and patient concerns about their appearance with worn corsets (2, 16, 64, 65).

#### 2.8.4 Surgical treatment

an indication for surgical treatment is the Cobb angle> 40-50° and the rapid progression of the curvature (51).

### 2.9. Prognosis

Risk factors for the progression of scoliosis are: female gender, time remaining until full skeletal growth, scoliotic curvature region and magnitude of scoliotic curvature progression being greater at the peak of skeletal growth in both infants and adolescents (16). Females have a higher degree of scoliotic curvature progression than males in the 8: 1 ratio (14). Age is the weakest and most variable indicator to determine full growth therefore bone maturity is taken as an indicator. Menarche status in women helps determine the peak of growth. With the onset of the menstrual cycle begins and the stage with the fastest bone growth lasting approximately 12 months, the growth of the spine ends 2 years later. The Risser sign provides reliable information on bone growth: if Risser 0 or 1 the risk of progression is 60-70% (16, 2). The low-value Risser sign, the early Tanner stage, and menarche indicate the progression of scoliosis (57). Closure of the triradiate cartilage of the acetabulum and ossifying centers in the bones of the hand (distal part of the ulna and radius, palm bones) is also a radiographic sign which quite accurately determines the time of peak growth. The ossifying centers of the hand are compared to the atlas of Greulich and Pyle. The shape and region of the scoliotic curvature predicts progression quite well: curves with apex above T12 progress more than lumbar scoliotic curves. The greater the magnitude of the curvature at first the greater the risk of progression. The magnitude of the scoliotic curvature at early 20-29° has a risk of progression above 68% (16, 2). Recently, the ScoliScore AIS prognostic genetic test is used, which the higher the values, the greater the possibility of progression (55).

## III. Purpose of the paper

Given that idiopathic adolescent scoliosis is quite heterogeneous and complex, it affects an age group of 10-18 years important for society, they are diseases that manifest secretly and have a long and tedious treatment procedure, special attention should be paid. The purpose of this paper is to describe the characteristics, etiology, epidemiology, pathogenesis, classification, clinical manifestations, diagnosis, prognosis and non-operative treatment of adolescent idiopathic scoliosis in the period 2018 September-2019 March. Through the paper we will try to present: the number of cases with juvenile idiopathic scoliosis, the number of cases divided by gender, the number of cases based on clinical manifestations, the prevalence of the disease, scoliotic curves according to the level of localization along the spine, association with kyphosis, types of scoliotic curves according to Cobb angle, form of non-operative treatment and number of cases for relevant forms of treatment.

## IV. Material and methods

The study is based on quantitative retrospective research, in the period September 2018-March 2019 (7 months) at the University Hospital and Clinical Service of Kosovo (SHSKUK) - Orthopedic Clinic, which analyzed data on 250 patients with juvenile idiopathic scoliosis in the age group 10 -18 years old with Cobb angle 10° who came from different parts of Kosovo. The data were obtained and used from the archive of the orthopedic specialist outpatient clinic system at the HUCSK, with special permission from the Office for Personal Data Protection and the Ethical-Professional Committee of the HUCSK. Data on the number of children aged 10-18 are taken from the latest data from the Kosovo Agency of Statistics. The collected data were analyzed, compared and presented in tabular form and through graphs. The number of patients aged 10-18 years with adolescent idiopathic scoliosis was calculated, analyzed and processed based on gender, symptomatology, which were reported to the orthopedic clinic, the region which included scoliosis, association with kyphosis, angle of Cobb and the required form of non-operative treatment. Microsoft Excel is used to extract, analyze and compare data.

## V. Results

The study included 250 patients with adolescent idiopathic scoliosis with Cobb angle ≥10° treated on an outpatient basis in orthopedic specialist clinics-SHSKUK, including the period September 2018-March 2019 (7 months). Out of a total of 10097 patients examined in orthopedic specialist clinics, 626 patients or expressed in percentage 6.19% of them were diagnosed with scoliosis. Out of 626 patients with scoliosis 250 or expressed in percentage 39.93% of them are diagnosed with adolescent idiopathic scoliosis which belong to the age group 10-18 years. Based on these data one in 40 patients reported to orthopedic specialist clinics is with adolescent idiopathic scoliosis. The prevalence of adolescent idiopathic scoliosis based on the data obtained is 1.40%. Meanwhile, the prevalence based on gender is 0.93% for women and 0.46% for men. By gender 167 patients out of the total number were female or expressed in percentage 66.8% and 83 patients or 33.2% were male. The ratio of females to males goes in favor of females 2.01: 1.

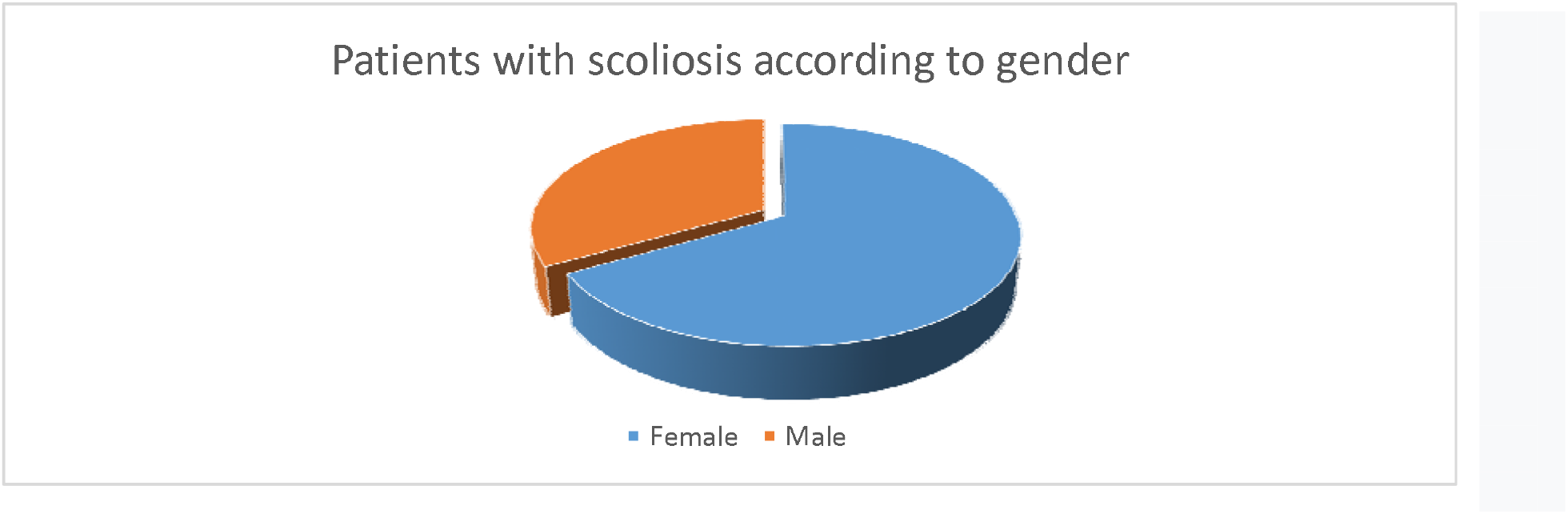

The reason that patients came to orthopedic clinics in 18.4% of cases out of the total number was back pain, while the rest are reported due to significant deformity of the spine, accidental finding during diagnostic procedures for other diseases and due to guidance by the doctor during screening done in primary schools. All of these patients with adolescent idiopathic scoliosis have tested positive for Adams.

Based on the region of the spine involved in scoliosis we have the following data: no case with cervical scoliosis, 7 cases with cervico-thoracal scoliosis (2.8%) from them 6 (2.4%) female patients and 1 (0.4%) male patients, 61 (24.4%) cases with thoracal scoliosis from them 36 (14.4%) female patients and 25 (10.0%) male patients, 152 (60.8%) cases with thoraco-lumbal scoliosis from them 107 (42.8%) female patients and 45 (18.0%) male patients, 29 (11.6%) cases with lumbal scoliosis from them 18 (7.2 %) female patients and 11 (4.4%) male patients, 1 (0.4%) case with lumbo-sacral scoliosis and it is a male patient. According to the data, there is a larger number of cases involving the thoraco-lumbar region from adolescent idiopathic scoliosis with 60.8% while the cervical region is not affected at all. And in all regions involved predominance of cases by female patients is observed.

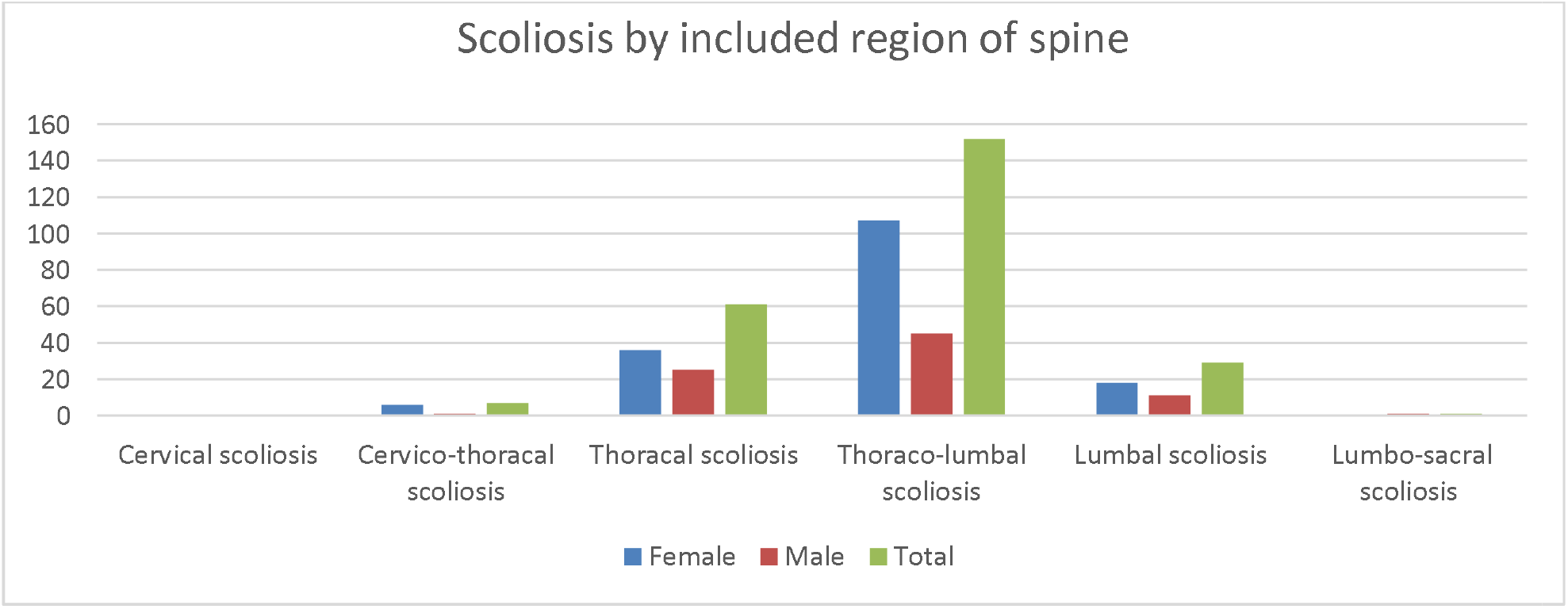

Adolescent idiopathic scoliosis includes 90 cases or 36% of cases with only one primary scoliotic curvature and 160 cases or 64% with 2 scoliotic curvatures: primary and secondary.

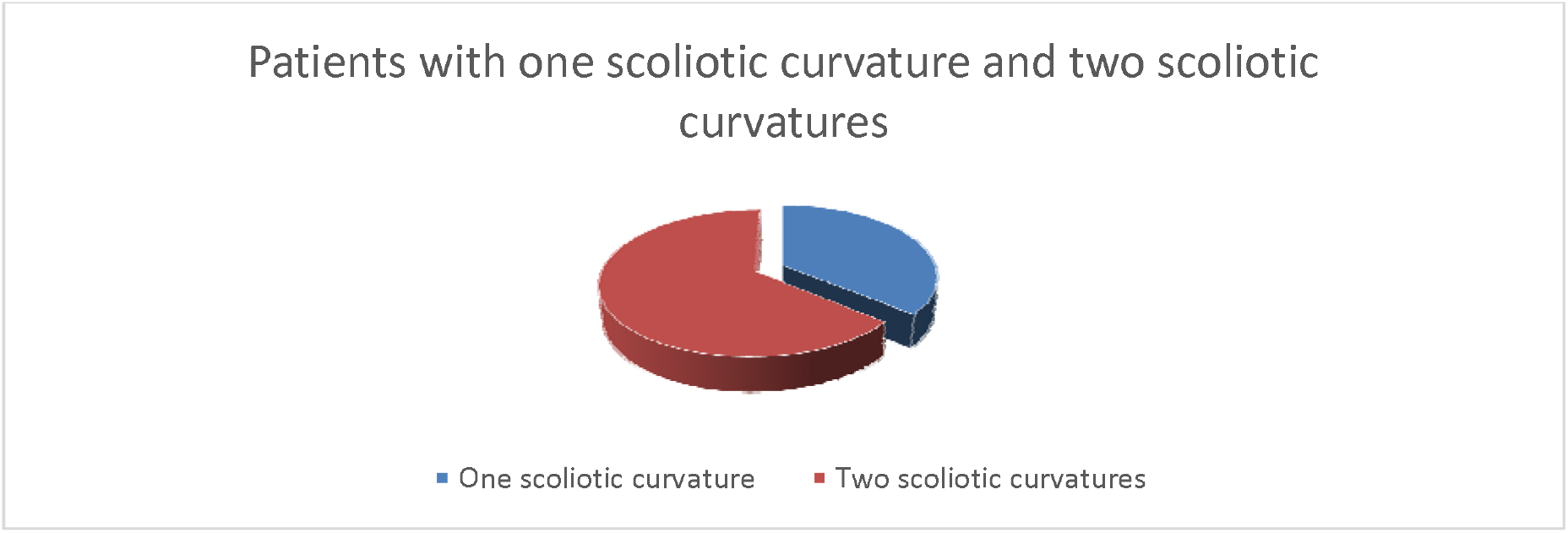

There are also cases of juvenile idiopathic scoliosis accompanied by kyphosis where there are a total of 21 cases or 8.4% of the total number.There was no case of cervical scoliosis associated with kyphosis, 3 (14.2%) cases with cervico-thoracal scoliosis associated with kyphosis and all of them are female, 8 (38.09%) cases with thoracal scoliosis associated with kyphosis from them 4 (19.04%) are female and 4 (19.04%) are male, 10 (47.61%) cases with thoraco-lumbal scoliosis associated with kyphosis from them 8 (38.09%) are female and 2 (9.52%) are male, and no cases where reported from lumbal and lumbo-sacral scoliosis associated with kyphosis. Assessing the total number of scoliosis based on the region involved, it is noticed that kyphosis is more associated with adolescent idiopathic scoliosis of the cervico-thoracic region with 42.8%, then of the thoracic region with 13.1% and finally of the thoraco-lumbar region with 6.51% from the total number of cases. Kyphosis based on these data does not accompany scoliosis of the cervical, lumbar and lumbo-sacral region.

Out of 250 cases collected only 108 patients or 43.2% of them have Cobb angle determined and written in medical reports.

| Cobb angle | Female | Percentage | Male | Percentage | Total | Percentage |
| --- | --- | --- | --- | --- | --- | --- |
| Mild curvature 10-30° | 56 | 51.85% | 39 | 36.11% | 95 | 87.96% |
| Moderate curvature 31-50° | 7 | 6.48% | 3 | 2.77% | 10 | 9.25% |
| Severe curvature > 50° | 3 | 2.77% | 0 |  | 3 | 2.77% |

The scoliotic curvature based on the Cobb angle is divided in this way and from the data the following results are obtained: mild curvature 10-30° were 95 (87.96%) patients from them 56 (51.85%) were female and 39 (36.11%) were male, moderate curvature 31-50° were 10 (9.25%) patients from them 7 (6.48%) were female and 3 (2.77%) were male, severe curvature > 50° were 3 (2.77%) cases and all of them female.There is a larger number of cases with mild scoliosis, ie 0-30° and easily manageable. It is also observed that with the increase of the Cobb angle the female / male ratio increases in favor of the female. While in light curves the female / male ratio is 1.4: 1, in moderate curves the female / male ratio is 2.3: 1, in heavy curves this ratio increases significantly more> 3: 1 in favor of females based on the total number data. Cobb angle cases> 50° were cases associated with kyphosis and involving two regions of the spine: thoracic and thoraco-lumbar.

These cases have undergone non-operative treatment according to the protocol for scoliosis

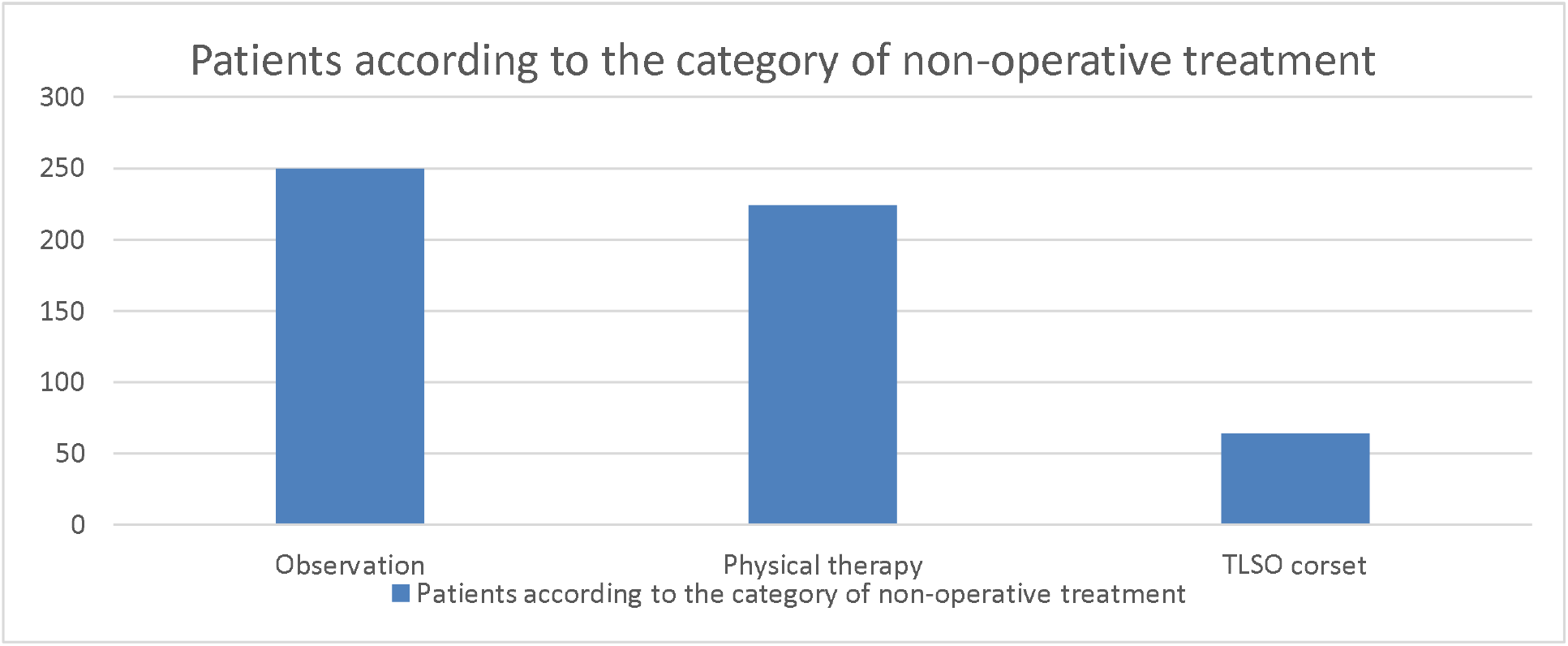

There are 250 patients with adolescent idiopathic scoliosis who are observed, of which 19 patients or 7.6% in addition to observation also receive pain therapy or Ibuprofen. According to this value, out of 46 patients who are referred to orthopedic specialist clinics with back pain, 19 patients or 41.3% receive pain therapy. Meanwhile, according to the use of one, two or three categories of non-operative treatment from the total number of cases, the following data were obtained: 26 patients or 10.4% of the total number were only kept under observation, 224 patients or 89.6% were observed and underwent physical therapy and 64 patients or 25.6% were observed, underwent physical therapy and wore the TLSO corset. Observation includes regular RTG checks of the AP and LL spine every 2 or 3 months as needed, physical therapy includes regular exercises to strengthen the back muscles and the applied corset was of the TLSO type and it is decided to stop the progression of the curvature scoliotic and reducing it, observation in these patients includes RTG of the AP and LL spine with corsets worn and without it. Patients who have been kept under observation and physical therapy belong to the group with Cobb angle 10-20°, while patients with Cobb angle> 20° in addition to observation and physical therapy have also worn corsets.

## VI. Discussion

The study included 250 patients aged 10-18 years diagnosed with juvenile idiopathic scoliosis with Cobb angle ≥10° treated on an outpatient basis in orthopedic outpatient clinics-SHSKUK, including the period September 2018-March 2019 (7 monthly). The prevalence of juvenile idiopathic scoliosis out of the total number of cases is 1.40%, an approximate value of the prevalence of a study conducted in Singapore in 1985. This study included 110,744 children aged 11-12 years and 16-17 years of age. who were examined and a prevalence value of 1.0% was obtained (67). While another research conducted in Greece in 1997 included in the research 89,901 children aged 9-14 years and gained a prevalence of 1.7% which also supports the prevalence of 1.40% (68). Of the total number of cases, women are more affected by adolescent idiopathic scoliosis than men with a ratio of 2.01: 1. These results are roughly supported by a 2017 study conducted in Indonesia, who based on data obtained from screening children aged 9-16 in schools won a female-to-female ratio of 1: 4.7 (66). The survey conducted in Singapore has a female / male ratio of 2: 1 (67) while another survey conducted in Germany in 2007 speaks of a 1.5-1 ratio in favor of females (69).

In orthopedic specialist clinics patients with juvenile idiopathic scoliosis have been reported due to deformity, accidental finding in the diagnosis of other diseases as well as due to doctor’s instruction from screening done in primary schools, only 18.4% of patients out of the total number of cases have been reported due to back pain. A 1997 study conducted by Ramirez with associates included 2442 patients with adolescent idiopathic scoliosis and during research on the prevalence of back pain and its association with other pathologies found that 23% of these patients have back pain in disease incidence (43) this value is approximately 18.4% of patients found in the total number of cases. Regarding the type of scoliosis there were no patients with cervical or sacral scoliosis. There are 7 patients or 2.8% with cervico-thoracic scoliosis, 61 patients or 24.4% with thoracic scoliosis, 152 patients or 60.8% with thoraco-lumbar scoliosis, 29 patients or 11.6% with lumbar scoliosis and 1 patient with lumbo-sacral scoliosis. Scoliosis with 1 main scoliotic curve makes up 36% of the total number and with 2 curves, one compensator 64%. Another study conducted in Singapore in 2005 shows the thoraco-lumbar curves as the most common with 40.1% of cases, then thoracic curves with 33.3% of cases as single curves, in third place are double and triple curves other than thoraco-lumbar curves with 18.7% of cases and finally lumbar curves with 7.9% of cases (70). This study supports the validity of these results.

Among other things, the association of adolescent idiopathic scoliosis with kyphosis was analyzed, where out of the total number, 21 cases or 8.4% of them were found. 3 or 42.8% cases were found associated with cervico-thoracic scoliosis out of the total number, 8 or 13.1% with thoracic scoliosis and 10 or 6.51% with thoracolumbar scoliosis. According to a 2019 study by Tang et al., there is talk of a slightly higher incidence of kyphosis in the cervical region in patients with adolescent idiopathic scoliosis than in patients who have no other pathology (70).

Patients diagnosed with juvenile idiopathic scoliosis undergo treatment according to the scoliosis protocol. From the total number of cases that all 250 patients were observed by means of RTG of the AP and LL spine and with re-examinations which depending on the need were done every 2 or 3 months, 224 patients or 89.6% of the total number of cases were instructed for physical therapy to strengthen the back muscles, relieve pain and thus stop further progression of scoliosis and 64 patients or 25.6% were instructed to wear TLSO corset to stop the progression of scoliotic curvature and reduce it to the maximum possible. Patients are usually referred to more than one form of non-operative treatment of scoliotic curvature with the exception of 26 patients or 10.4% of cases for which it is only reasonable to observe. Of the 250 patients who were observed, ibuprofen pain therapy was given to 19 patients or 7.6% of the total number of 46 patients or 18.4% who had back pain. While 224 patients or 89.6% of the total number were observed and underwent physical therapy and 64 patients or 25.6% of the total number were observed, underwent physical therapy and wore TLSO type corsets.

Of the 250 patients diagnosed with adolescent idiopathic scoliosis only 108 patients or 43.2% of the total number possess the Cobb angle in medical reports. From this number according to Cobb angle curves we have 95 patients or 87.9% of the total number with curve with Cobb angle 10-30°, 10 patients or 9.2% with curve with Cobb angle 31-50 ° and 3 patients or 2.7% with Cobb angle> 50°. While in light curves the female / male ratio is 1.4: 1, in moderate curves the female / male ratio is 2.3: 1, in heavy curves this ratio increases significantly more> 3: 1 in favor of females based on the total number data. Many studies like the one in Singapore in 1985 (67), the study conducted in Korea in 2011 (72) report for greater Cobb angle in girls than in boys resulting in the fact that scoliotic curvature in girls progresses to higher rate than in boys. For patients with a Cobb angle greater than 30° the ratio increases more than 10: 1. Patients with juvenile idiopathic scoliosis with Cobb angle ≥ 20° from general data were instructed to wear corsets, underwent physical therapy, and were observed. According to a 2012 study by Schlenzka and Yrjönen, the indications for corsets in patients with adolescent idiopathic scoliosis are the Cobb angle> 25° and with progression ≥ 5° so that the corset is effective, otherwise for smaller angles it is sufficient physical therapy and observation (62).

## VII. Conclusion

During the period September 2018-March 2019 in the specialist orthopedic clinics-HUCSK were diagnosed and treated 250 cases of idiopathic adolescent scoliosis of the age group 10-18 years with Cobb angle 10° ≥. By gender, women are affected more than men in a 2: 1 ratio. The reasons for reporting to orthopedic specialist clinics by patients were deformity, accidental detection of the disease during the diagnosis of other diseases, instruction by doctors during screening in schools and in 18.4% of cases back pain. The most common form of adolescent idiopathic scoliosis is the form which affects the thoraco-lumbar region with 60.8% of cases. 21 cases out of the total number are associated with kyphosis including scoliotic curves of the cervico-thoracic, thoracic and thoracic region. There are 89.7% of cases belonging to the group with slight curves according to the Cobb angle and the female / male ratio is increasing in favor of the female with the increase of the Cobb angle. Patients underwent one or more forms of non-operative treatment: observation 10.4% of cases, observation and physical therapy 89.6% of cases and observation, treatment and wearing of TLSO corset 25.6% of cases. Cases involving all three forms of non-operative treatment are with Cobb angle ≥20°. Based on these data it is recommended to continue screening in children’s schools so that scoliosis is noticed earlier and treated earlier. It is recommended to prepare medical staff to perform the Adams test and other scoliotic measurements so that the process is more comprehensive. Such steps will reduce the number of patients with scoliosis and the costs of treating it.

## Data Availability

The gathered data are shown below.

https://drive.google.com/file/d/1_9vGshgie-FRtZFNeDR_EMd7Z3hwtM4o/view?usp=sharing

